# Associations between reasons for vaping and current vaping and smoking status: Evidence from a UK based cohort

**DOI:** 10.1101/19006007

**Authors:** Jasmine N. Khouja, Amy E. Taylor, Marcus R. Munafò

**Affiliations:** MRC Integrative Epidemiology Unit at the University of Bristol, United Kingdom; Bristol Medical School: Population Health Sciences, University of Bristol, United Kingdom; UK Centre for Tobacco and Alcohol Studies, School of Psychological Science, University of Bristol, United Kingdom; NIHR Biomedical Research Centre at the University Hospitals Bristol NHS Foundation Trust and the University of Bristol, United Kingdom

**Keywords:** ALSPAC, e-cigarettes, smoking

## Abstract

**Background:** This study aimed to discover which young adults vape, the reasons given for vaping, and which reasons for vaping are associated with continued vaping/smoking.

**Methods:** In a UK cohort of 3,994 young adults, we explored the association of retrospectively-recalled reasons for vaping by 23 years with vaping/smoking status at 24 years. Using logistic regression, we assessed the association with vaping behaviour among ever vapers who had ever smoked (n=668), and with smoking behaviour among individuals who regularly smoked prior to vaping (n=412).

**Results:** Vaping to quit smoking was associated with higher likelihood of vaping (odds ratio [OR] = 3.51, 95% confidence interval [95%CI] = 2.29 to 5.38), but lower likelihood of smoking at 24 years (OR = 0.50, 95%CI = 0.32 to 0.78). Vaping to cut down smoking was associated with higher likelihood of vaping (OR = 2.90, 95%CI = 1.87 to 4.50) and smoking at 24 years (OR = 1.62, 95%CI = 1.02 to 2.58). Vaping out of curiosity was associated with lower likelihood of vaping at 24 years (OR = 0.41, 95%CI = 0.26 to 0.63) but higher likelihood of smoking at 24 years (OR = 1.66, 95%CI = 1.04 to 2.65).

**Conclusions:** Intention to quit smoking appears important for young adults to stop smoking using e-cigarettes; vaping to cut down is associated with continued smoking, but smoking to quit is associated with discontinued smoking. Vaping out of curiosity is less likely to lead to a change in smoking/vaping behaviour (i.e., current smokers continue to smoke).

## 1. INTRODUCTION

Of an estimated 3.6 million vapers in Great Britain, over half now consider themselves ex-smokers (Action on Smoking and Health, 2019). Evidence suggests e-cigarettes are less harmful than cigarettes (Public Health England, 2015) and can effectively aid smoking cessation (Hajek et al., 2019), but some smokers have not tried e-cigarettes, and not all who have tried them have successfully quit smoking (Hartmann-Boyce et al., 2016; Zhu et al., 2017). Given their popularity, it is important to know which individuals use e-cigarettes, why, and whether different reasons for use are associated with continued use of e-cigarettes and smoking cessation.

One study in the US showed that vapers are more likely to be male and have a lower income (Levy et al., 2017). Additionally, smokers and ex-smokers who had quit in the last 3 years were more likely to have used e-cigarettes than those who had not, but a lower percentage of current and regular vapers were current smokers compared to ever vapers. In the UK, current and non-current vapers differ in socio-economic status, number of cigarettes smoked per day and past year quit attempts (Brown et al., 2014). However, there is limited research exploring whether these differences are observable among young adults in the UK.

Among adults (18+ years) in Great Britain, the primary reasons for vaping are related to smoking cessation (Action on Smoking and Health, 2019). However, evidence from the US suggests that young adults vape primarily out of curiosity (Kong et al., 2015) or because their friends/family vape (Tsai et al., 2018). In a study of South Korean adolescents, the most common reason for vaping among infrequent users was out of curiosity, but the most common reasons for frequent vaping were to quit smoking and vape indoors (Lee et al., 2017). Vaping to quit smoking was also associated with continued vaping among US middle and high school students (Bold et al., 2016). Although some studies have explored associations between reasons for vaping and continuation/discontinuation of vaping and smoking (Bold et al., 2016; Nicksic et al., 2019; Saddleson et al., 2016; Yong et al., 2019), there is limited evidence from the UK. Young adults in their mid-twenties are a relatively understudied subgroup in research on reasons for vaping; most research focusses on school children, students or older adults.

We aimed to explore the characteristics of young adults who are vaping or have previously vaped, and whether different reasons for vaping are associated with later vaping and smoking among ever smokers and ever vapers. Specifically, we sought to investigate whether different retrospectively recalled reasons for vaping by 23 years are associated with vaping and/or smoking one year later among a UK cohort of young adults.

## 2. METHODS

### 2.1 Study Population

Young adults enrolled in the Avon Longitudinal Study of Parents and Children (ALSPAC) formed the study sample. The profile of the cohort has previously been described in two publications (Boyd et al., 2013; Fraser et al., 2013) and the phases of enrolment are described in more detail in the cohort profile update (Northstone et al., 2019). The total sample size for the cohort is 15,454 pregnancies, resulting in 15,589 foetuses (Figure 1). Please note that the study website contains details of all the data that is available through a fully searchable data dictionary and variable search tool (http://www.bristol.ac.uk/alspac/researchers/our-data/). ALSPAC study data from 22 years onwards were collected and managed using REDCap electronic data capture tools hosted at the University of Bristol (Harris et al., 2009).

**Figure 1.**
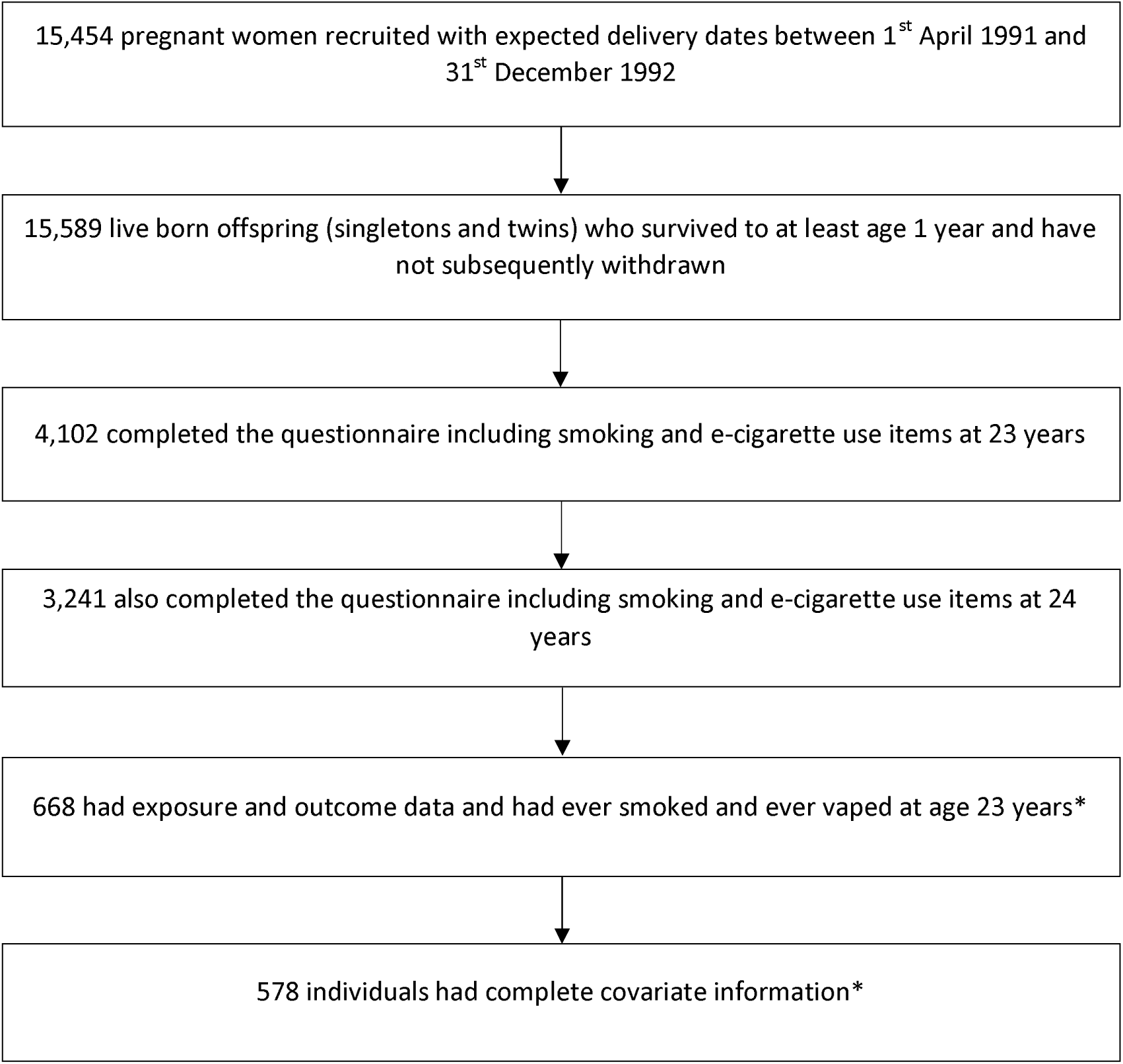
Flow chart depicting the process of data inclusion in the analysis of the associations between reasons for vaping at 23 years and vaping and smoking at 24 years. *Figures shown for ever vapers and ever smokers, for ever vapers and prior smokers n = 412 and n = 360 respectively.

The questionnaires on vaping and smoking at 23 years of age were completed by 4,102 children (here on in described as young adults) and 3,241 of these also completed the questionnaires at 24 years. Figure 1 displays the recruitment process.

Ethics approval for the study was obtained from the ALSPAC Ethics and Law Committee and the Local Research Ethics Committees. Informed consent for the use of data collected via questionnaires and clinics was obtained from participants following the recommendations of the ALSPAC Ethics and Law Committee at the time.

### 2.2 Measures

#### 2.2.1 Participant characteristics

We measured a range of behavioural and lifestyle factors to compare the characteristics of: (i) ever vapers (vaped at least once) versus never vapers; and (ii) former vapers (vaped at least once but reported not currently vaping at 23 years) versus self-reported current vapers. Maternal smoking in pregnancy was recorded at 18 weeks gestation. Body mass index (BMI) was measured in clinic at 17 years. Measures of risk-taking were taken at 20 years (alcohol, tobacco, marijuana and other drugs and gambling). Education/employment status and parenthood status (i.e., had become a parent or not) were measured at 21 years. Mental health factors, measured at 21 years, were anxiety (GAD-7) and low mood in the past 4 weeks. Factors relating to vaping and smoking were measured at 23 and 24 years.

#### 2.2.2 Exposure

To determine whether questionnaire respondents had ever vaped, they were asked “Have you ever used/vaped an electronic cigarette (e-cigarette) or other vaping device?” To determine whether they had ever smoked, they were asked “Have you ever smoked a whole cigarette (including roll-ups)?” To determine whether respondents were regular smokers immediately prior to vaping (i.e., prior smokers), they were asked “Did you smoke tobacco regularly just before you started using electronic cigarettes/vaping devices?” In a multiple-choice question at 23 years, respondents who had ever vaped before were asked “what are/were your reasons for using electronic cigarettes/vaping devices?” and instructed to cross all answers that applied. Seven response options were available: “To help me quit smoking”, “To help me cut down on the number of cigarettes I smoke”, “To help me with cravings in situations where I cannot smoke e.g. travel, indoors”, “Pleasure”, “Curiosity”, “Friends use them” and “Other”. The “Other” option was rarely selected so was not included in this analysis.

#### 2.2.3 Outcome

At 24 years, e-cigarette outcome data was collected via questionnaire on current vaping. Current vaping was self-reported by the respondent in response to the question “Do you currently use/vape e-cigarettes or other vaping devices?” To determine smoking status at 24 years, the young adults were asked “Have you smoked any cigarettes in the past 30 days?”

#### 2.2.4 Potential confounders

Various demographic factors were measured which have been shown to impact the likelihood of vaping and smoking and could influence the reason given for vaping (Hiscock et al., 2012; Jamal et al., 2016). Sex was recorded at birth. Parental socioeconomic position (SEP; assessed via occupational status) was recorded at 18 weeks gestation. Ethnicity was recorded at 32 weeks gestation.

#### 2.2.5 Further information

Further information is provided in the Supplementary Material. This includes information regarding the questionnaires and variables used.

### 2.3 Statistical Analysis

#### 2.3.1 Differences in participant characteristics

Differences between never and ever vapers as well as former and current vapers were assessed using a χ^2^ test or t-test.

#### 2.3.2 Reasons for e-cigarette use and smoking/vaping behaviour

In a series of logistic regressions, we explored the association between retrospectively reported reasons for vaping by 23 years and vaping and smoking continuation at 24 years. Each of the six reasons for vaping were analysed individually as a binary variable (indicated/not indicated as a reason for use). Vaping and smoking status were treated as binary variables (i.e., current vaper versus not current vaper and smoker versus not current smoker). Firstly, we explored the association between reasons for vaping (retrospectively reported at 23 years) and current vaping at 24 among ever vapers and ever smokers at 23 years. Secondly, we explored the association between reasons for vaping (retrospectively reported at 23 years) and current smoking at 24 among ever vapers at 23 who were regular smokers just prior to first e-cigarette use. These regressions were analysed with and without adjustment for demographic factors (sex, ethnicity, parental SEP, and age in months at 23-year questionnaire).

#### 2.3.3 Supplementary analysis

As we cannot determine whether individuals were dual using products in the main analysis, we further explored the association between reasons for vaping and later vaping and smoking status using multinomial logistic regression. Vaping and smoking status was categorised into four groups: current smoker (smoking but not currently vaping), dual user (currently vaping and smoking), current vaper (vaping but not currently smoking), or neither user (not currently vaping nor smoking). Analyses were adjusted for demographic factors (sex, ethnicity, parental SEP, and age). Analyses were restricted to ever vapers who had 1) ever smoked at 23 years and 2) regularly smoked just prior to vaping.

#### 2.3.4 Multiple imputation

Of the young adults who completed the questionnaire on vaping at 23 years, 62% stated they had ever vaped, ever smoked and had complete data including all covariates (22% were missing outcome data, 14% were missing covariate data). We used multiple imputation, a recommended method to account for missing data (Sterne et al., 2009), to increase the sample size available for analysis and minimise bias due to attrition. Data was imputed for missing covariate information; all those included in the analysis had complete exposure and outcome data. Adjusted analyses were repeated using multiply imputed data. The multiple imputation by chained equations procedure was completed using the ICE package in Stata 15.1 which created 100 datasets with 20 cycles. Data were imputed for the 668 young adults who completed the questionnaires at 23 and 24 years and responded that they had ever smoked and ever used an e-cigarette. Further details of the imputation, including the auxiliary variables used, can be found in Supplemental Table 1.

**Table 1.**
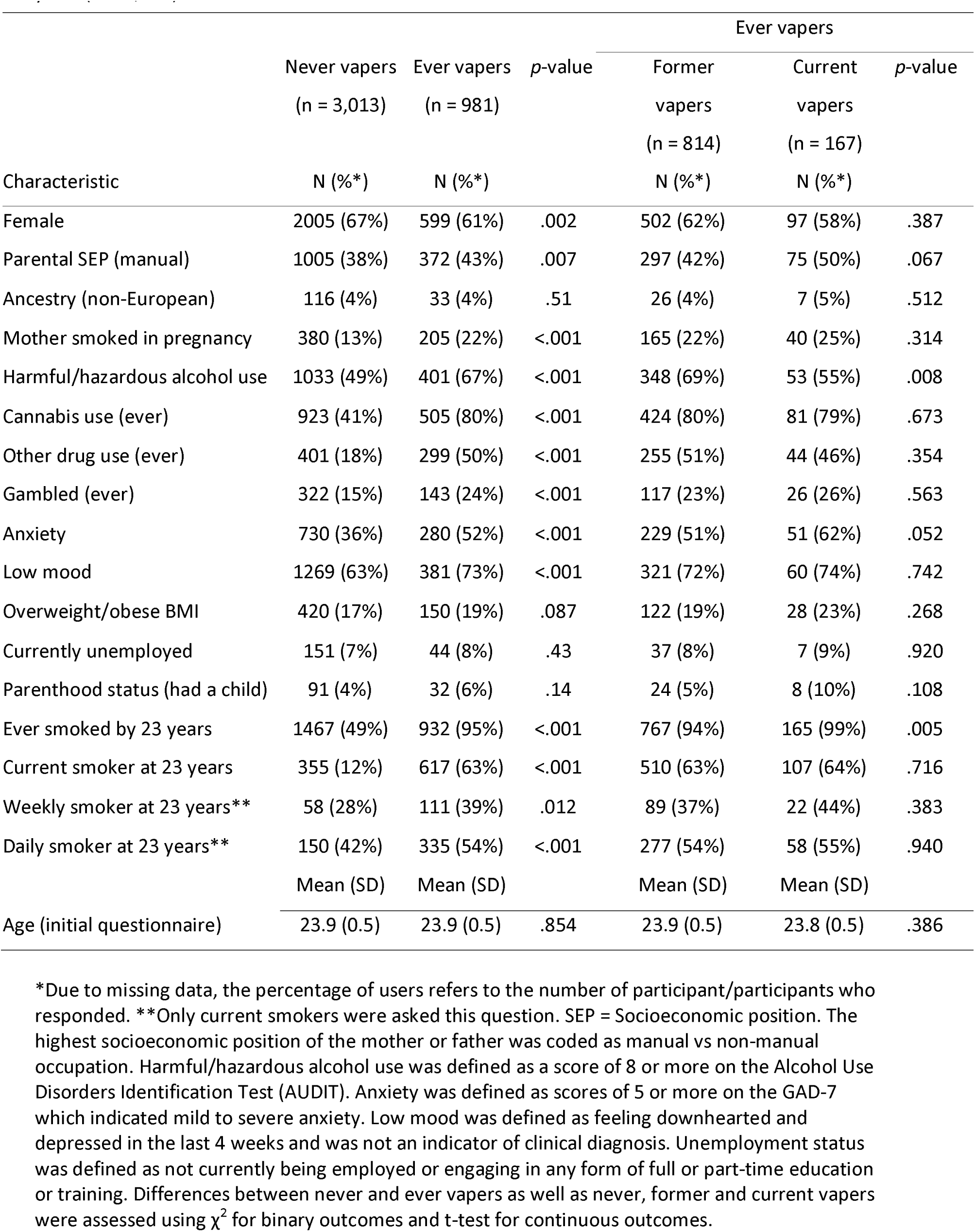
Characteristics of the study population for never, former and current e-cigarette users at 23 years (N = 3,994).

#### 2.3.5 Minimum detectable effect

A power calculation revealed that we had sufficient power (90%) to detect a minimum odds ratio of 1.51. Details on the calculation of the minimum detectable effect are provided in the Supplementary Materials.

## 3. RESULTS

### 3.1 Characteristics of Vapers at Age 23

The characteristics of the young adults are described in Table 1, grouped into never vapers (n = 3,013), and ever vapers (n = 981); ever vapers were also grouped into former vapers (n = 814) and current vapers (n = 167). A higher percentage of ever vapers were male, were of lower parental SEP at birth, had a mother who smoked in pregnancy, and engaged in other potentially addictive or harmful behaviours (harmful or hazardous drinking, ever drug use, gambling). Ever vapers were also more likely to report anxiety or low mood, smoking by the age of 20 or 23 years, and be current, weekly or daily smokers than never vapers at 23 years. Vaping among never smokers was rare; 5% of ALSPAC participants who had ever vaped had never smoked, and <1% of ever vapers defined themselves as current vapers who had never smoked at 23 years. There were no clear differences in ethnicity, BMI, unemployment or parenthood between these groups. On average, participants were 23 years old across all groups at the initial questionnaire. The age at which young adults first vaped was similar among current and former vapers (median = 22 years of age). There were few clear differences between former and current vapers. Current vapers were more likely to have lower parental SEP at birth, report anxiety and have smoked by the age of 23 years but were less likely to be hazardous or harmful alcohol users than former vapers.

### 3.2 Reasons for Vaping

Participants’ reasons for vaping by 23 years are shown in Table 2. Current vapers were more likely than former vapers to vape for all reasons except ‘out of curiosity’, the most popular reason given for vaping (51%). Most young adults (56%) selected one reason for vaping (Supplemental Table 2).

**Table 2.** Reasons for e-cigarette use among former and current vapers by 23 years (N = 981).

|  | Former<br>vapers<br>(n = 814) | Current<br>vapers<br>(n = 167) |  | Full sample<br>(n = 981) |
| --- | --- | --- | --- | --- |
| Reasons for vaping by 23 | N (%) | N (%) | *p*-value | N (%) |
| Vaped to quit smoking | 222 (27%) | 113 (68%) | <0.001 | 335 (34%) |
| Vaped to cut down number of cigarettes smoked | 161 (20%) | 68 (41%) | <0.001 | 229 (23%) |
| Vaped to help with cravings in situations when unable to smoke | 64 (8%) | 41 (25%) | <0.001 | 72 (7%) |
| Vaped for pleasure | 119 (15%) | 54 (32%) | <0.001 | 173 (18%) |
| Vaped out of curiosity | 469 (58%) | 32 (19%) | <0.001 | 501 (51%) |
| Vaped because friends used them | 186 (23%) | 22 (13%) | 0.005 | 208 (21%) |

### 3.3 Reasons for Vaping and Smoking/Vaping Behaviour

Due to small numbers of never smokers who had tried vaping at 23 years (n = 47), analyses were restricted to ever-smokers who had ever vaped. The study sample consisted of 668 young adults who had completed both questionnaires and had ever vaped and smoked by 23 years. 412 of these young adults were regular smokers immediately prior to vaping. The median time between 23- and 24-year questionnaire completion was 12 months. The age at which the young adults first vaped ranged from 17 to 24 years (SD = 1; median = 22) and on average occurred 2 years before questionnaire completion. Supplemental Table 3 displays vaping characteristics for vapers at 23 years. At 24 years, 49% of the young adults (n = 330) were current smokers. Dual use (n = 62) and current vaping (n = 47) were less common. A substantial proportion of the young adults were neither users at 24 years (n = 229).

The unadjusted and unimputed adjusted results of the logistic regressions are shown in Supplemental Tables 4 and 5. The imputed adjusted results are shown in Table 3. The results were consistent, with all associations in the same direction with a similar magnitude.

**Table 3.** Associations between reasons for vaping by 23 years and current vaping at 24 years among ever vapers and ever smokers and current smoking at 24 years among ever vapers and prior smokers.

| Reason for vaping by 23 years | Vaping at 24 years (n=668) |  |  |  | Smoking at 24 years* (n=412) |  |  |  |
| --- | --- | --- | --- | --- | --- | --- | --- | --- |
|  | N (%) | OR | 95% CI | p-value | N (%) | OR | 95% CI | p-value |
| To quit smoking | 228 (34%) | 3.51 | 2.29, 5.38 | <.001 | 217 (53%) | 0.50 | 0.32, 0.78 | .002 |
| To cut down | 166 (25%) | 2.90 | 1.87, 4.50 | <.001 | 158 (38%) | 1.62 | 1.02, 2.58 | .041 |
| To curb cravings | 75 (11%) | 4.35 | 2.57, 7.37 | <.001 | 70 (17%) | 0.84 | 0.47, 1.49 | .553 |
| Pleasure | 118 (18%) | 3.22 | 2.01, 5.15 | <.001 | 57 (14%) | 0.88 | 0.47, 1.65 | .685 |
| Curiosity | 351 (53%) | 0.41 | 0.26, 0.63 | <.001 | 158 (38%) | 1.66 | 1.04, 2.65 | .035 |
| Friends used them | 153 (23%) | 0.63 | 0.37, 1.09 | .10 | 71 (17%) | 1.78 | 0.95, 3.36 | .073 |

The analyses were run on multiply imputed data for individuals who ever smoked and ever vaped. Both analyses adjusted for demographic factors (sex, ethnicity, socioeconomic position, and age in months at 23-year questionnaire). \*This analysis was restricted to those who had reported that they smoked regularly just before they started vaping. OR = Odds ratio. Note: OR = Odds ratio.

Vaping to quit smoking by 23 years was associated with higher likelihood of continuing to vape at 24 years (odds ratio [OR] = 3.51, 95% confidence interval [95%CI] = 2.29 to 5.38, *p <* 0.001). Amongst individuals smoking just prior to e-cigarette use, vaping to quit smoking was associated with lower likelihood of continuing to smoke at 24 years (OR = 0.50, 95%CI = 0.32 to 0.78, *p* = 0.002) and higher likelihood of being a dual user, current vaper, or neither user compared to being a current smoker at 24 years (Supplemental Table 6).

Vaping to cut down the number of cigarettes smoked was associated with higher likelihood of continuing to vape at 24 years (OR = 2.90, 95%CI = 1.87 to 4.50, *p <* 0.001). Amongst individuals smoking just prior to first e-cigarette use, vaping to cut down was associated with higher likelihood of continuing to smoke at 24 years (OR = 1.62, 95%CI = 1.02 to 2.58, *p =* 0.041) and higher likelihood of being a dual user compared to being a current smoker at 24 years (Supplemental Table 6).

Vaping to curb cravings for cigarettes (OR = 4.35, 95%CI = 2.57 to 7.37, *p <* 0.001) and for pleasure (OR = 3.22, 95%CI = 2.01 to 5.15, *p <* 0.001) were also associated with higher likelihood of continuing to vape at 24 years. Both were associated with higher likelihood of being a dual user compared to a current smoker at 24 years (Supplemental Table 6) among those who were smokers just prior to first using e-cigarettes. Among the full sample, vaping for pleasure was also associated with higher likelihood of being a current vaper and neither user compared to being a current smoker at 24 years (Supplemental Table 6).

Vaping out of curiosity was associated with lower likelihood of continuing to vape (OR = 0.41, 95%CI = 0.26 to 0.63) and a higher likelihood of continuing to smoke at 24 years (OR = 1.66, 95%CI = 1.04 to 2.65, *p* = 0.035). Among the full sample, vaping out of curiosity was associated with lower likelihood of being a dual user and higher likelihood of being a neither user compared to being a current smoker at 24 years (Supplemental Table 6). However, for those who were smokers just prior to first e-cigarette use, vaping out of curiosity was only associated with lower likelihood of being a current vaper compared to a current smoker at 24 years (Supplemental Table 6).

Vaping because friends vaped was associated with higher likelihood of continuing to smoke at 24 years (OR = 1.78, 95%CI = 0.95 to 3.36, *p* = 0.073 among prior smokers). Among those who were smokers just prior to first e-cigarette use, vaping because friends vaped was associated with lower likelihood of being a neither user compared to a current smoker at 24 years (Supplemental Table 6).

## 4. DISCUSSION

The results indicate that there are substantial differences in characteristics between young adults who have never and ever vaped at 23 years. Vaping out of curiosity and to quit smoking were common reasons for vaping among young adults in this UK-based sample. Five reasons for vaping by 23 years included in this study were strongly associated with continued vaping at 24 years; vaping to quit smoking, cut down, curb cravings, and for pleasure were associated with higher likelihood of continued vaping whereas vaping out of curiosity was associated with lower likelihood of continued vaping. Four reasons for vaping by 23 years were associated with continued smoking at 24 years; vaping to quit smoking was associated with lower likelihood of continued smoking whereas vaping to cut down, out of curiosity and because friends vaped were associated with higher likelihood of continued smoking.

Similar to previous findings (Brown et al., 2014), we found that vapers were more likely to have lower parental SEP at birth than never vapers. Vapers were also more likely to report risk taking behaviours (e.g., drug use, gambling) and poorer mental health. Smoking has previously been associated with similar characteristics (Hiscock et al., 2012; Jamal et al., 2016; Lai et al., 2000; Minichino et al., 2013). This could indicate a common liability for both behaviours (i.e., the same factors increase the likelihood of engaging in both behaviours) which could explain why not all young adults who vape go on to quit smoking. Alternatively, smoking could mediate the relationship between risk taking and vaping; most vapers in this cohort smoked prior to vaping and some started vaping to quit smoking. In line with previous findings (Levy et al., 2017), vapers were more likely to be ever, weekly or daily smokers at 23 years, and vaping among never smokers was rare. Even though 52% of the sample did not provide a smoking-related reason for vaping and we cannot determine order of product use for all participants, it is unlikely that vaping led to smoking in this cohort; only 1% of the young adults stated they were not regularly smoking *just before*, but started smoking regularly *after* using an e-cigarette.

Our results imply that vaping to quit smoking may facilitate young adults’ quit attempts as those who regularly smoked prior to vaping for this reason were less likely to continue to smoke at 24 years and were more likely to be neither users than current smokers. This is in line with the growing amount of evidence that vaping can facilitate smoking cessation (Hajek et al., 2019; Hartmann-Boyce et al., 2016; Polosa et al., 2015; Public Health England, 2015). Our findings support previous findings among middle and high school students (Bold et al., 2016) and college students in the US (Saddleson et al., 2016) which found that vaping to quit smoking was associated with continued vaping. For some young adults, vaping to quit smoking could encourage the continued use of e-cigarettes rather than quitting nicotine products entirely. If young adults aim to be nicotine free, they may need support to quit vaping or reduce the levels of nicotine in their e-liquid once they have successfully quit smoking.

Those who were regular smokers just prior to starting to vape and who vaped to cut down were more likely to still be smokers at 24 years than those who did not vape to cut down. Similar to previous findings (Bold et al., 2016) they were also more likely to continue vaping (i.e., be dual users) which suggests that these individuals were not encouraged to quit smoking. There are several potential explanations for this. First, these users may not actually intend to quit smoking, and intention may be necessary for e-cigarettes to act as an effective smoking cessation tool. Second, the comparison group includes those who vaped with the sole intention to quit smoking which could potentially mask a decreased likelihood of continuing smoking among those who vaped to cut down. However, adjusting for use to quit smoking did not substantially change the results (OR = 1.62, 95%CI = 1.01 to 1.60). Third, recall bias may be an issue; those who were unable to quit smoking may report that they only intended to cut down. Fourth, these users may still be in the process of quitting and may eventually quit entirely.

Consistent with research conducted in the US (Bold et al., 2016; Kong et al., 2015), we found that curiosity is the most common reason for vaping among young adults, and these users were less likely to continue vaping at 24 years and were more likely to be neither users than current smokers. As the main sample are ever smokers who have ever vaped, this may highlight a group of ‘experimenters’ who try both vaping and smoking but do not continue either behaviour. Those who were regular smokers just prior to vaping out of curiosity were more likely to continue smoking and less likely to continue vaping, which implies that smokers who vape out of curiosity do not want to or are unable to quit smoking compared to those who vape for other reasons. The evidence suggests that vaping out of curiosity does not lead to a change in smoking/vaping status; ever smokers were unlikely to become current vapers after trying e-cigarettes out of curiosity and were more likely to be neither users than current smokers, but those who were regular smokers when they tried e-cigarettes out of curiosity were likely to remain current smokers at 24 years.

Although there was some missing covariate data which could have introduced selection bias, we used multiple imputation methods to address this. However, the study was limited by the measures included, particularly the exposure; the reasons for vaping were self-reported retrospectively. This measure could suffer from recall bias; young adults may be less likely to state that they vaped to quit smoking if they were unsuccessful in their attempt. Also, the reasons for vaping chosen were not exclusive for each young adult due to the multiple-choice format of this question. Furthermore, open questions and qualitative analysis may have uncovered other potential reasons for vaping which were omitted from this analysis. An ‘other’ option provided in the questionnaire where young adults could provide open answer responses but this was rarely selected (4% of the sample) or selected without another reason being specified (2%) so was not included in the analysis. Additionally, the answer options that were provided may have primed a response in the young adults which was biased towards smoking cessation as three were smoking related.

The timing of this cohort study may impact the generalisability of the findings. E-cigarettes are a relatively new product compared to cigarettes; consequently, cigarettes were available to this cohort of young adults for a considerable period of their adolescence before e-cigarettes became widely available in 2007. In 2007, the study sample were roughly 17 years old and cigarette initiation peaks at around 15-16 years of age (Marcon et al., 2018), so it is likely that these young adults experimented with cigarettes prior to being exposed to e-cigarettes, young adults today are being exposed to both e-cigarettes and cigarettes during adolescence and this may have an impact on their reasons for vaping as well as their current vaping and smoking status. Although it would be interesting to observe the association between reasons for vaping and later vaping and smoking status among those who were never smokers when they first tried vaping, we are unable to accurately identify these individuals with the current available data. It would also be interesting to observe the associations among never smokers at 23 years but there are too few individuals who had vaped but never smoked at 23 to report any meaningful analysis.

### 4.1 Conclusions

Vaping out of curiosity does not appear to change young adults smoking and vaping status. Vaping to quit smoking is associated with continued vaping and discontinued smoking. In contrast, vaping to cut down was associated with continued smoking. This implies that intention to quit smoking may be necessary for young adults to effectively stop smoking using e-cigarettes. Further research is needed to explore the potential implications of these findings using stronger causal inference methods (e.g., randomising young adults to cut down or quit smoking to explore the impact of intention on smoking cessation using e-cigarettes).

## Data Availability

The ALSPAC study website contains details of all the data that is available through a fully searchable data dictionary and variable search tool (http://www.bristol.ac.uk/alspac/researchers/our-data/). Data access is restricted but can be requested (http://www.bristol.ac.uk/alspac/researchers/access/).

http://www.bristol.ac.uk/alspac/researchers/access/

## Acknowledgements

We are extremely grateful to all the families who took part in this study, the midwives for their help in recruiting them, and the whole ALSPAC team, which includes interviewers, computer and laboratory technicians, clerical workers, research scientists, volunteers, managers, receptionists and nurses.

## Contributors

All authors (JNK, AER and MRM) contributed to the design of the study and JNK collated and analysed the data. JNK produced the first of the article which was edited and approved by AER and MRM.

